# The moderating impact of the genetic predisposition to smoking behaviour on the response to tobacco excise taxes

**DOI:** 10.1101/2020.12.02.20242388

**Authors:** Eric A.W. Slob, Cornelius A. Rietveld

## Abstract

Tobacco consumption is one of the leading causes of preventable death. While some public policies have been effective in reducing the smoking prevalence in the United States, high tobacco excise taxes do not appear to deter all individuals from starting smoking nor to affect the smoking intensity of all those who do smoke. Here, we analyse whether someone’s genetic predisposition to smoking may explain why individuals smoke despite high tobacco excise taxes. For this purpose, we interact polygenic risk scores for smoking behaviour with state-level excise tax rates on tobacco. Our analyses exploiting longitudinal data (1992-2016) from the US Health and Retirement Study show that someone’s genetic propensity to smoking moderates the effect of tobacco excise taxes on smoking behaviour along the extensive margin (smoking vs. not smoking) and the intensive margin (the amount of tobacco consumed). That is, when tobacco excise taxes are relatively low, those with a high genetic predisposition to smoking are more likely (i) to smoke, and (ii) to smoke a relatively high number of cigarettes per day. In our sample, we do not find a significant interaction effect on smoking cessation.

## 1. Introduction

Tobacco use is the leading preventable cause of death in the world, causing over 7 millions deaths per year (World Health Organization, 2017). In the United States, over 480,000 deaths per year are attributable to smoking (US Department of Health and Human services, 2014). Tobacco use has been shown to be quite addictive and hence, quitting is often a tough battle characterized by heavy withdrawal symptoms (Benowitz, 2008). As a prime instrument to influence smoking behaviour, governments impose excise taxes on tobacco. Over the past 50 years, the median price of cigarettes has increased from 0.30$ per pack up to 5.70$ (US Department of Health and Human services, 2014). In the same period, consumption per capita decreased from 4, 000 to about 1, 000 cigarettes per year. Although this decrease cannot entirely be explained by the increase in tobacco excise taxes in this period, because for example public awareness about the detrimental effects of smoking also increased in this period, there is convincing evidence about the effectiveness of raising tobacco excise taxes for reducing smoking (Chaloupka & Warner, 2000; Institute of Medicine, 2007; MacLean et al., 2016). However, the decrease in smoking consumption has stalled in the past 20 years (Orzechowski & Walker, 2016).

Tobacco excise taxes are similar for each member of a society, and a possible explanation for the stabilizing smoking prevalence in the US may be that for some individuals it is more difficult than for others to stop smoking. For example, studies have shown that demand elasticities for tobacco differ between males and females (Yen, 2005) and across ethnicities (Kandel et al., 2004). Moreover, behavioural preferences such as risk aversion (Anderson & Mellor, 2008; Barsky et al., 1997) and someone’s health status influence smoking behaviour (Clark & Etilé, 2002; Jones, 1994; Lahiri & Song, 2000). There is also clear evidence that heavy smokers react differently to tobacco excise taxes than less heavy smokers (Nesson, 2017), although the precise mechanism explaining these elasticity differences is not known. In the present study, we analyze whether someone’s genetic predisposition to smoking moderates the response to tobacco excise taxes.

Several studies have shown that the heritability of smoking behaviour ranges between 31-60 % (Bidwell et al., 2016), indicating that genes explain a considerable proportion of the variation in smoking in a population possibly through their effect on nicotine dependency. It has also been shown that environmental circumstances such as state policies impact the heritability of smoking: The heritability of smoking is lower in states with relatively high excise taxes on tobacco and in states with greater controls on cigarette advertising and vending machines (Boardman, 2009). Recent large-scale genetic association studies have found more than 500 genetic variants underlying the heritable variation in smoking behaviour (Erzurumluoglu et al., 2020; Liu et al., 2019). These genetic variants are expressed in biological systems that affect reward processing and addiction (Liu et al., 2019). Fletcher (2012) shows that individuals carrying one of these genetic variants respond differently to excise tobacco taxes than those not carrying this genetic variant. Hence, such a gene-environment interaction may explain why certain individuals smoke and others do not when tobacco excise taxes are high.

However, a follow-up study by Fontana (2015) using the same genetic variant shows that Fletcher’s gene-environment interaction was a spurious association that should be explained by the effects of population stratification. Population stratification entails an association between genetic subpopulations in a population and environmental conditions, such as cultural and social norms (Rietveld et al., 2014). Besides, recent studies have shown that the predictive power of individual genetic variants is limited, often below 0.02% for behavioural outcomes including smoking (Chabris et al., 2015). Hence, low statistical power may be another reason for why Fontana (2015) could not replicate the results of Fletcher (2012).

To deal with the limited predictive power of genetic variants, methods have been developed to combine multiple genetic variants into a composite genetic measure. The most often adopted approach is the construction of so-called polygenic risk scores (PGSs) (Dudbridge, 2013). To construct a PGS, all genetic variants in a sample are summed up in a weighted fashion in which each weight is proportional to the strength of the association between the genetic variant and an outcome variable as estimated in a genome-wide association study (GWAS) (International Schizophrenia Consortium, 2009). For example, a recent study shows that polygenic scores currently explain about 4% of the variance in smoking behaviour (Liu et al., 2019). A polygenic score not only makes one well powered for out of sample prediction, but also enables more powerful gene-environment interaction analysis. However, by using polygenic scores, Fontana (2015) shows that the interaction between someone’s genetic predisposition (as captured by the polygenic scores for educational attainment and smoking intensity) and tobacco excise taxes is insignificant in a model explaining the intensity of tobacco consumption

The present study adopts the same approach to study gene-environment interaction in smoking behaviour as Fontana (2015), but goes beyond the study by Fontana in four ways. First, whereas Fontana uses polygenic scores for educational attainment and smoking intensity, we use a set of polygenic scores more directly related to smoking behaviour. That is, we use polygenic scores specifically constructed for smoking initiation, smoking intensity and smoking cessation. Second, we use polygenic scores that are more predictive for smoking behaviour. The predictive power of polygenic scores is strongly dependent on the sample size of the GWAS on which results the polygenic scores are based (Dudbridge, 2013). We use polygenic scores based on the recent results of the *GWAS & Sequencing Consortium of Alcohol and Nicotine use* (GSCAN) (Liu et al., 2019) which were obtained in a sample of *N* > 1,1 million individuals (∼15 x more individuals than in the smoking GWAS that Fontana used to construct polygenic scores). Third, through the inclusion of additionally genotyped individuals (∼12,000 vs. ∼8,500) as well as non-genetic data from the three most recent waves of data collection from the US Health and Retirement Study, our analyses are better powered. As such, we have higher chances of estimating significant interaction effects. Fourth, whereas Fontana focusses on the intensity of smoking only, we analyse the intensive margin (smoking vs. not smoking), the extensive margin (number of cigarettes smoked per day), and smoking cessation (smoking continuation vs. smoking cessation). As such, we provide a more complete analysis of smoking behaviour.

Establishing a G × E interaction is often complicated by the fact that individuals with a certain genetic predisposition may self-select into certain environments (Jencks, 1980). In this study, we overcome bias from such a gene-environment correlation by exploiting exogenous variation in the level of tobacco excise rates across states and years. As such, this study provides the first robust evidence of the existence of a gene-environment (G × E) interaction influencing smoking behaviour. Our results suggest that individuals with a higher genetic propensity for smoking respond more strongly to a change in excise taxes compared to individuals with a lower genetic propensity. When tobacco excise taxes are relatively low, those with a high genetic predisposition to smoking are more likely to smoke. If smoking, those with a high genetic predisposition to smoking consume a relatively high number of cigarettes per day when tobacco excise taxes are low. In our sample, we do not find a significant interaction effect on smoking cessation. Thus, in line with Boardman (2009), our study shows that genetic effects are more pronounced when environments are less restrictive (i.e., when tobacco excise taxes are relatively low).

Our study mainly contributes to two streams of literature. First, we enrich the literature analysing smoking behaviour and responses to tobacco excise taxes (Anderson & Mellor, 2008; Barsky et al., 1997; Boardman, 2009; Clark & Etilé, 2002; Jones, 1994; Kandel et al., 2004; Lahiri & Song, 2000; Nesson, 2017; Yen, 2005) by overcoming limitations of earlier studies analysing G × E interactions on smoking (Fletcher, 2012; Fontana, 2015). Second, we contribute to an emerging literature on gene-environment interactions (G × E) exploiting *exogenous* variations in environments that addresses how the environment moderates the effect of genetic variants, and vice versa (Barcellos et al., 2018; Conley & Rauscher, 2013; Schmitz & Conley, 2016; Pereira et al., 2020; Schmitz & Conley, 2017). These studies stress that the analysis of exogenous variation in environments is key to overcome bias from gene-environment correlation when estimating G × E interactions.

The remainder of this paper is organized as follows. In section 2, we introduce the genetic and non-genetic data we draw on in our study. Section 3 describes our methodological approach. In section 4 we present our main results and section 5 provides a discussion of our results and concludes.

## 2. Data Description

The data used in this study are derived from the US Health and Retirement Study (HRS) (Juster & Suzman, 1995). The HRS is a longitudinal survey consisting of approximately 20,000 individuals who were surveyed biennially since 1992. The respondents in the survey are a representative sample of Americans over age 50 and their spouses. The HRS aims to analyze the health and behaviour of individuals approaching or just after retirement. Therefore, the dataset includes information about for example work status, pension plans, income, health insurance, physical health and functioning, cognitive functioning, and health behaviours including drinking and smoking (for an overview, see Karp (2007)). From 2006 onwards, the HRS started to collect genetic data from their respondents. In the present study, we exploit data collected in the waves from 1992 up to 2016 (13 waves in total) which have been harmonized by the RAND Corporation (RAND HRS Longitudinal File 2016 V2).

### 2.1. Smoking behaviour

The main outcomes in the present study are three measures of smoking behaviour. The first (binary) variable is based on the question ‘Do you smoke cigarettes now?’, and equals 1 if an individual is currently smoking and 0 otherwise. The second (continuous) variable is the response to the question ‘About how many cigarettes or packs do you usually smoke in a day now?’. This question is only asked to individuals who are currently smoking, and it is set to 0 in case an individual does not smoke. The third (binary) variable measures smoking cessation, and is constructed using our first smoking variable. It equals 1 if an individual smokes at time (interview wave) *t* — 1 and t, and 0 if an individual smokes at time *t* — 1 but not at time *t*.

### 2.2. State-level excise tobacco taxes

The Tax Burden on Tobacco dataset (Orzechowski & Walker, 2016) provides us information about the tax levied by the state on each purchased pack of cigarettes (based on the state and federal tax in each year). We converted these nominal prices to real prices using the Consumer Price Index from the US Bureau of Labor Statistics (US Bureau of Labor Statistics, 2018), using 1991 as the base year. These data were merged with the HRS data, based on confidential data about the state the HRS respondent currently lives in. As the HRS contains biennial survey data, we use the tax levied in the year prior to each survey. For consistency with prior studies and to facilitate the interpretation of effects as proportional changes in consumption, the tax levels are logarithmically transformed (Adda & Cornaglia, 2006; Fletcher, 2012).

### 2.3. Polygenic scores

Polygenic scores are used to analyse whether the response to tobacco excise taxes is moderated by someone’s genetic predisposition to smoking. Most genetic differences across individuals in a population can be attributed to single nucleotide polymorphisms (SNPs). A SNP is a location in the DNA strand at which two different nucleotides can be present in the population. For each SNP, an individual’s genotype is coded as a 0, 1 or 2, depending on the number of reference nucleotides present. Individuals who inherited the same nucleotide from each parent are called homozygous for that SNP (and have genotype 0 or 2), while individuals who inherited different nucleotides are called heterozygous (and have genotype 1). Polygenic scores reflect the combined additive influence of SNPs on a particular outcome.

To construct a polygenic score, SNPs are summed up in a weighted fashion. The weights reflect the strength of the relationship between a SNP and the outcome of interest, as estimated in a GWAS. In a GWAS, for each SNP the following model is estimated:

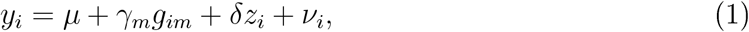

where *y*_*i*_ is the outcome of interest for individual *i, μ* is an intercept, *γ*_*m*_ is the additive effect of SNP *g*_*im*_, *z*_*i*_ is a vector of control variables (e.g., sex and age),*ν*_*i*_ and is the residual. Using the effect size estimates *γ*_*m*_ from (1), the polygenic score is constructed as:

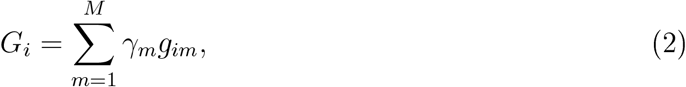

where *G*_*i*_ represents the value of the polygenic score for individual *i, M* is the total number of SNPs included in the construction of the polygenic score, *γ*_*m*_ is the additive effect size of SNP *m* taken as estimated in the GWAS and *g*_*im*_ is the genotype of individual *i* at locus *m* (measured as 0, 1 or 2).

The HRS provides polygenic scores for public distribution based on the results of several recently conducted large-scale GWASs (Ware et al., 2018). In this study, we use three polygenic score to measure someone’s genetic predisposition to smoking behaviour. The first polygenic score is based on the results of a GWAS on *smoking initiation (ever smoked vs. never smoked)*, and measures someone’s genetic predisposition to start smoking. The second polygenic score is based on the results of a GWAS with the *number of cigarettes smoked per day* as dependent variable. As such, the second score reflects someone’s genetic predisposition to heavy smoking. Lastly, we have a polygenic score that is based on the results of a GWAS on *smoking cessation (currently smoking vs. smoking formerly)*. This captures someone’s genetic predisposition to being able to stop smoking after having started smoking. Hence, the first polygenic score reflects the genetic predisposition for smoking on the intensive margin, the second one reflects the genetic predisposition for smoking on the extensive margin, and the third one for smoking cessation.

The weights for constructing the polygenic scores come from the GWASs conducted by the GSCAN consortium (Liu et al., 2019). In total, approximately 1.4 million SNPs were used to construct the polygenic scores (Ware et al., 2018). We use the polygenic scores constructed for individuals of European ancestry in the sample, because the GSCAN analyses were also restricted to individuals of this ancestry. To facilitate an easy interpretation of the results, the polygenic scores are standardized such that they have mean 0 and a standard deviation of 1 in the analysis sample. Higher values reflect a higher genetic predisposition to smoking behaviour. For smoking cessation, we reverse coded the polygenic score such that a higher score reflects a higher chance to remain smoking.

### 2.4. Covariates

For comparability purposes, the choice of individual level control variables is based on the studies by Fletcher (2012) and Fontana (2015). We include an individual’s gender as a covariate, to control for differences between males and females. Furthermore, we include an individual’s birth year to account for possible age specific differences in smoking behaviour and we add birth year squared to account for possible non-linearities in age effects. We account for the socio-economic status of the respondent by including individual income (as imputed by the RAND Corporation, see Hurd et al. (2016), in real terms using 1992 as base year) and years of education (self-reported by participants) in the model.

Although Fontana controls for the change in health status in his models, we abstain from it because of possible endogeneity issues (Lahiri & Song, 2000). Compared to Fletcher’s model, we do not control for race/ethnicity because we restrict our sample to individuals of recent European ancestry. This is a commonly used restriction in genetic studies and also recommended by the genotyping center, as this restriction pre-empts possible bias from population stratification (Weir, 2012). That is, correlations between allele frequencies and environmental factors across subpopulations in the overall HRS sample. To deal with more subtle forms of population stratification in the analysis sample, we include the first 10 genetic principal components of the genetic relationship matrix as control variables (Ware et al., 2018). The genetic relationship matrix includes pairwise genetic relationships between individuals in the sample as estimated using SNPs. It has been shown that the inclusion of principal components solves the problem of subtle population stratification adequately in the HRS (Rietveld et al., 2014).

Finally, we include both state dummies and wave dummies to account for differences across states and over time.

## 3. Methods

To test for the presence of an effect of the interaction between someone’s genetic predisposition to smoking behaviour and tobacco excise taxes on smoking outcomes, we use a moderation framework for each of our three outcome variables. The baseline regression for explaining whether an individual is currently smoking is given by:

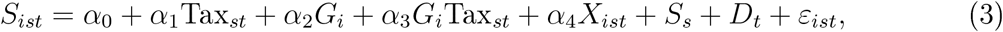

where *S*_*ist*_ is a binary variable indicating whether individual *i* residing in state *s* in year *t* is currently smoking or not, Tax_*st*_ represents the cigarette tax in state *s* at year *t*, and *G*_*i*_ is the value of the polygenic score for individual *i. X*_*ist*_ represents the vector of individual-level control variables. The *α*’s represent the corresponding effect size estimates for these variables. The vectors *S*_*s*_ and *D*_*t*_ are vectors for state and year fixed effects. Lastly, *ε*_*ist*_ denotes the error term. Despite the binary nature of *S*_*ist*_, we estimate the model using linear regression to make the interpretation of the coefficient more straightforward and to avoid the difficulties surrounding the estimation of interaction effects in non-linear models.^1^

The response to taxes in terms of tobacco consumption is estimated by:

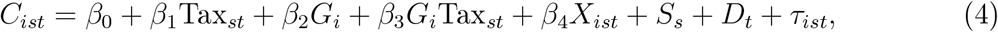

where *C*_*ist*_ denotes the number of cigarettes smoked per day by individual *i* at time *t* in state *s*. The other variables are the same as in equation 3. In this equation, the *β*’ s are the effect size estimates and *τ*_*ist*_ is the residual term. We estimate this model both in the full sample and in the subsample of smokers, because non-smokers are not likely to start smoking when tobacco excise taxes are increased.

Finally, we analyse smoking cessation using discrete-time survival models. Allison (Allison, 1982) shows that such survival models can be operationalized by using regression models for binary dependent variables. Therefore, we perform a binary logistic regression to explain the binary variable for smoking cessation. Importantly, this model can deal with right-censored observations, such as individuals who are still smoking in 2016. This model can be written as

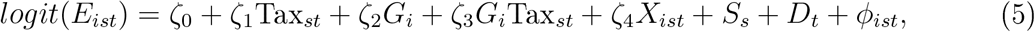

where *E*_*ist*_ denotes whether individual *i* in state *s* stopped smoking between time *t* — 1 and time *t*. In this regression, the ζ ‘s are the effect size estimates and *ϕ*_*ist*_ is the residual term. In our models, we do not use clustered standard errors as recommended by Allison (Allison, 1982). As a result of the inclusion of wave dummies in the models, the hazard rate (the probability that an individual stops smoking at between time *t* — 1 and time *t* given that (s) he has not yet done so at *t* — 1) is assumed to be different in each of the 13 waves in the sample.

## 4. Results

Table 1 contains the descriptive statistics of the analysis sample. It contains information about the full sample and the subsample of current smokers. Time-invariant variables are constant over the waves of data collection, but time-variant variables can take different values over time. In the full sample, there are more females than males and the mean birth year is 1941. There are small differences between the full sample and the subsample of smokers with respect to birth year, years of education, income, and marital status. Not surprisingly, the means of the polygenic scores for smoking behaviour as well as the smoking prevalence and the average number of cigarettes smoked per day are relatively high in the subsample of current smokers. The means for smoking cessation are the same in the full sample and in the subsample, because this variable is only constructed for those that smoked in at least one of the interview waves. Figure 1 shows that there is a gradual increase of tobacco excise taxes over time (in real terms) and that there is considerable variation across states regarding the level of tobacco excise taxes imposed.

**Table 1:** Descriptive statistics analysis sample.

| **Time-invariant variables** | Full sample |  | Subsample of current smokers |  |
| --- | --- | --- | --- | --- |
|  | Mean | Std. Dev. | Mean | Std. Dev. |
| Female | 0.57 | 0.50 | 0.56 | 0.50 |
| Birth year | 1941 | 11.94 | 1944 | 10.47 |
| Years of education | 13.26 | 2.53 | 12.56 | 2.35 |
| $PGS_{\text{Smoking initiation}}$ | 0.000 | 1.000 | 0.25 | 1.01 |
| $PGS_{\text{Smoking intensity}}$ | 0.000 | 1.000 | 0.14 | 0.99 |
| $PGS_{\text{Smoking cessation}}$ | 0.000 | 1.000 | 0.11 | 0.96 |
| **Time-variant variables** | Mean | Std. Dev. | Mean | Std. Dev. |
| Currently smoking | 0.14 | 0.34 | 1.00 | 0.00 |
| Cigarettes smoked per day | 2.32 | 7.26 | 17.10 | 11.65 |
| Smoking cessation | 0.15 | 0.36 | 0.15 | 0.36 |
| Income ($\times \$1,000$) | 10.91 | 29.06 | 11.83 | 22.42 |
| Married | 0.65 | 0.48 | 0.61 | 0.49 |
| Individuals | 12,089 |  | 2,643 |  |
| Observations | 106,000 |  | 14,386 |  |

*Notes:* Std. Dev. = Standard deviation. After casewise deletion, the analysis sample comprises 12,058 individuals (from which 2,620 are current smokers in at least one interview wave).

**Figure 1:**
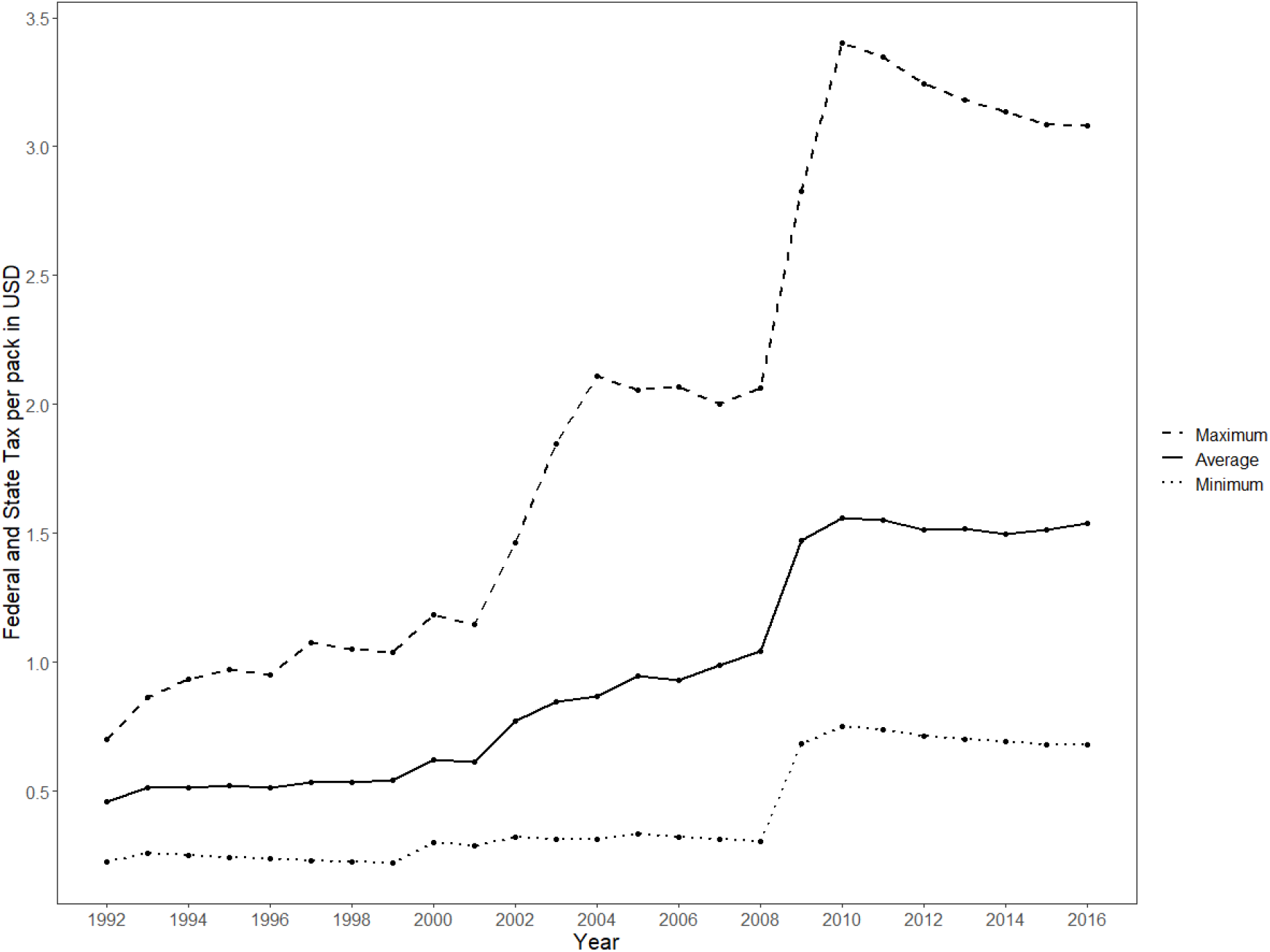
Real tobacco excise taxes levied per pack of 20 cigarettes.

Table 2 present the results of the model explaining whether an individual is currently smoking (the extensive margin). Column 1 shows that higher state-level tobacco excise taxes are negatively associated with the dependent variable^2^, and that the polygenic score for smoking initiation is positively associated with an individual’s current smoking status. Both these results are in line with expectations. In terms of effect sizes, an increase of excise taxes by 1% reduces the likelihood of smoking by about 7 percentage points, and an increase of one standard deviation in the polygenic risk score increases the chance of smoking by about 4 percentage points.

**Table 2:** Results of the regressions explaining an individual’s current smoking status.

|  | (1) | (2) | (3) |
| --- | --- | --- | --- |
| -Log(Tax) | 0.066***<br>(0.005) | 0.066***<br>(0.005) | 0.000<br>(0.005) |
| $PGS_{\text{Smoking initiation}}$ | 0.038***<br>(0.004) | 0.036***<br>(0.004) | 0.036***<br>(0.004) |
| $-\text{Log(Tax)} \times PGS_{\text{Smoking initiation}}$ | | 0.012***<br>(0.003) | 0.012***<br>(0.003) |
| Female | -0.027***<br>(0.005) | -0.027***<br>(0.005) | -0.031***<br>(0.005) |
| Birth year | 0.024<br>(0.077) | 0.022<br>(0.077) | -0.101<br>(0.074) |
| Birth year<sup>2</sup> | -0.000<br>(0.000) | -0.000<br>(0.000) | 0.000<br>(0.000) |
| Income ($\times \$1,000$) | -0.000***<br>(0.000) | -0.000***<br>(0.000) | -0.000***<br>(0.000) |
| Years of education | -0.016***<br>(0.001) | -0.016***<br>(0.001) | -0.016***<br>(0.001) |
| Married | -0.080***<br>(0.006) | -0.080***<br>(0.006) | -0.088***<br>(0.006) |
| State & Wave dummies | No | No | Yes |
| Observations | 105,959 | 105,959 | 105,959 |
| Individuals | 12,058 | 12,058 | 12,058 |
| $R^2$ | 0.0780 | 0.0783 | 0.0938 |

*Notes:* Standard errors in parentheses (clustered by state and individual); Coefficients for the constant term and the principal components are not reported, but available upon request from the authors; \* $p < 0.05$ , \*\* $p < 0.01$ , \*\*\* $p < 0.001$ .

In Column 2, the interaction term between the state-level tobacco excise taxes and the polygenic score for smoking initiation has been added to the model. This interaction term is significantly positive, indicating that low excise taxes on tobacco make those with a high genetic predisposition for smoking more likely to smoke. Column 3 shows that upon inclusion of state and wave fixed effects, the coefficient for tobacco excise taxes becomes insignificant. This change can be explained by the fact that tobacco taxes within a state increase in a monotonic fashion over time. These dynamics are absorbed by the state and wave dummies. However, the interaction term between the polygenic score and tobacco excise taxes remains statistically significant in Column 3.^3^

Table 3 presents the results of the regressions explaining someone’s smoking intensity (the intensive margin, in terms of cigarettes per day). In Column 1 (Full sample) and Column 4 (Subsample of current smokers), tobacco excise taxes are significantly negatively associated with the number of cigarettes smoked per day. In terms of effect sizes, an increase of excise taxes by 1% reduces cigarette consumption by 1.59 cigarettes per day in the full sample, and 3.48 cigarettes in the sample of current smokers. The polygenic score is again predictive of smoking behaviour (one standard deviation increase in the polygenic score leads to an increase in consumption of 0.36 cigarettes per day in the full sample and an increase of 1.08 in the subsample of current smokers). Column 2 show that the interaction effect is significantly positive in the full sample. This suggests that within the full sample, individuals with a high value for the polygenic score tend to consume more cigarettes than individuals with a low value for the polygenic score when tobacco excise tax decrease.^4^

**Table 3:** Results of the regressions explaining an individual’s smoking intensity.

|  | Full sample |  |  | Subsample of current smokers |  |  |
| --- | --- | --- | --- | --- | --- | --- |
|  | (1) | (2) | (3) | (4) | (5) | (6) |
| -Log(Tax) | 1.585***<br>(0.115) | 1.587***<br>(0.115) | 0.0239<br>(0.151) | 3.481***<br>(0.394) | 3.428***<br>(0.386) | 0.0661<br>(0.444) |
| $PGS_{\text{Smoking intensity}}$ | 0.358***<br>(0.0532) | 0.318***<br>(0.0482) | 0.319***<br>(0.0479) | 1.075***<br>(0.159) | 0.962***<br>(0.154) | 0.936***<br>(0.151) |
| $-\text{Log(Tax)} \times PGS_{\text{Smoking intensity}}$ | | 0.204**<br>(0.0591) | 0.205**<br>(0.0602) | | 0.376<br>(0.188) | 0.372*<br>(0.178) |
| Female | -0.901***<br>(0.0925) | -0.902***<br>(0.0929) | -0.980***<br>(0.0958) | -3.324***<br>(0.353) | -3.322***<br>(0.353) | -3.714***<br>(0.360) |
| Birth year | 1.937<br>(1.561) | 1.916<br>(1.569) | -0.958<br>(1.503) | 14.10*<br>(6.615) | 13.79*<br>(6.608) | 11.38<br>(6.452) |
| Birth year<sup>2</sup> | -0.000<br>(0.000) | -0.000<br>(0.000) | 0.000<br>(0.000) | -0.003*<br>(0.002) | -0.004*<br>(0.002) | -0.003<br>(0.002) |
| Income ($\times \$1,000$) | -0.005**<br>(0.002) | -0.005**<br>(0.002) | -0.009**<br>(0.003) | 0.000<br>(0.006) | 0.000<br>(0.006) | -0.014<br>(0.008) |
| Years of education | -0.337***<br>(0.0227) | -0.337***<br>(0.0228) | -0.321***<br>(0.0214) | -0.298**<br>(0.0865) | -0.298**<br>(0.0864) | -0.273**<br>(0.0846) |
| Married | -1.429***<br>(0.121) | -1.430***<br>(0.121) | -1.623***<br>(0.121) | -0.396<br>(0.291) | -0.391<br>(0.291) | -0.970**<br>(0.293) |
| State & Wave dummies | No | No | Yes | No | No | Yes |
| Observations | 105,930 | 105,930 | 105,930 | 14,254 | 14,254 | 14,254 |
| Individuals | 12,058 | 12,058 | 12,058 | 2,620 | 2,620 | 2,620 |
| $R^2$ | 0.0577 | 0.0580 | 0.0753 | 0.0654 | 0.0658 | 0.104 |

*Notes:* Standard errors in parentheses (clustered by state and individual); Coefficients for the constant term and the principal components are not reported, but available upon request from the authors; \* $p < 0.05$ , \*\* $p < 0.01$ , \*\*\* $p < 0.001$ .

When comparing the results in the full sample with those in the subsample of current smokers, we observe that the effect sizes are relatively large in the latter subsample. The estimates suggest that current smokers are reacting stronger to differences in taxes. This could be explained by the fact that smokers are able to reduce their smoking intensity, whereas in the full sample the non-smokers are not likely to change their smoking behaviour in response to increases in tobacco excise taxes (i.e., to start smoking). We observe that the interaction term in this subsample is not significant (Column 5). When adding state and wave dummies (Column 6), the interaction effect becomes significant, which strengthens our belief that the (borderline) insignificant finding in Column 5 is due to the smaller sample size of the subsample of current smokers.

In Table 4, the results for the logistic regression explaining smoking cessation are shown.^5^ Column 1 shows that tobacco excise taxes are significantly associated with smoking cessation. In terms of effect size, doubling excise taxes changes the odds of smoking cessation by 8.6%. We observe that the polygenic score is also predictive for cessation (a one standard deviation change in the polygenic score increases the odds of smoking cessation by 7.9%). The interaction term is insignificant, both in the model without state and wave dummies (Column 2) and in the model with these variables (Column 3).

**Table 4:**
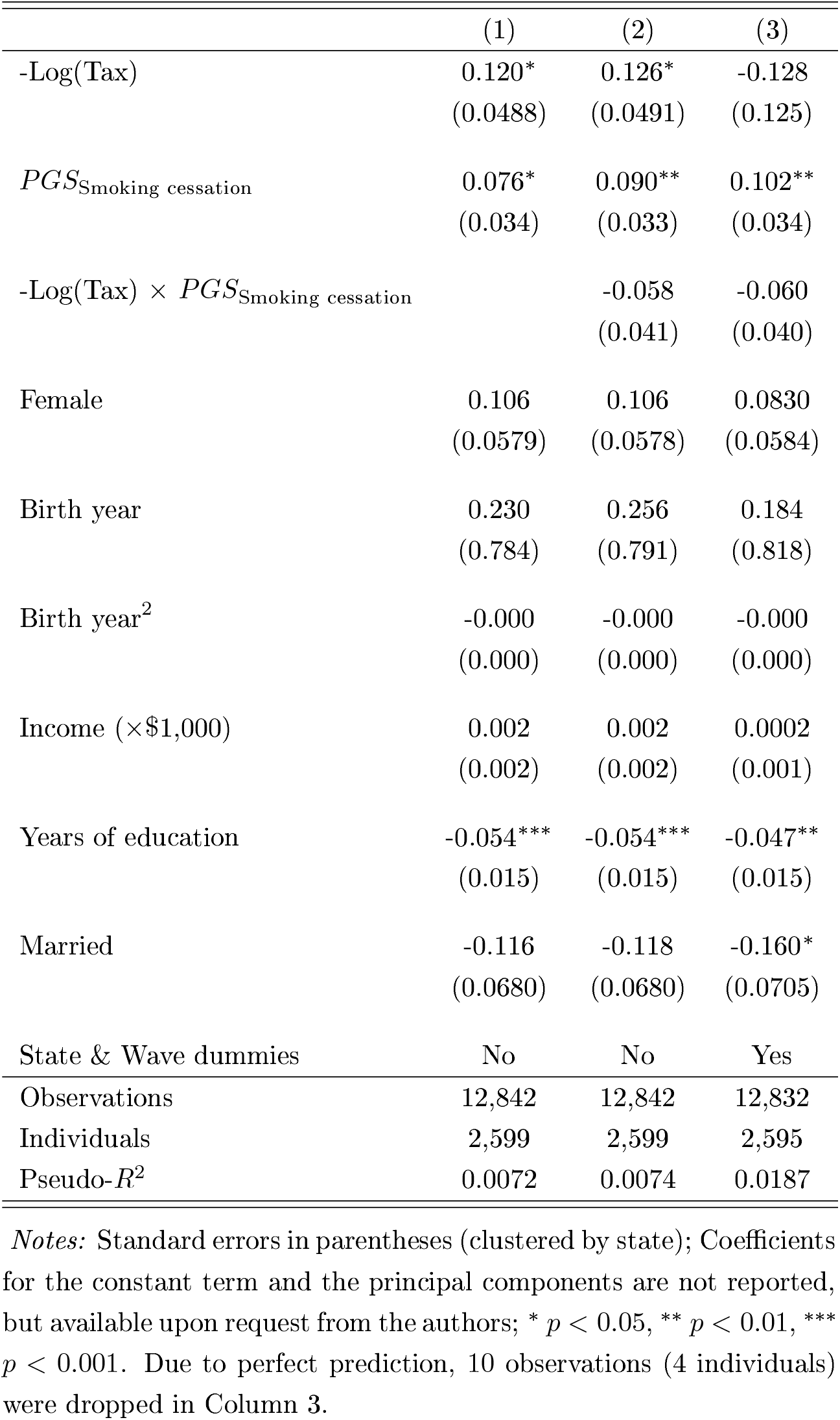
Results of the (logit) regressions explaining an individual’s decision to remain smoking.

In sum, the results show that the interaction between an individual’s genetic predisposition to smoking and state-level tobacco excise taxes significantly impacts whether someone smokes or not (the intensive margin) and the amount of tobaccco consumption (the extensive margin), but not smoking cessation.

## 5. Discussion and conclusion

The present study shows that someone’s genetic predisposition and tobacco taxes not only impact smoking behaviour additively, but also that someone’s genetic predisposition to smoking behaviour moderates the impact of tobacco excise taxes on tobacco consumption (both along the intensive and extensive margin). However, this interaction does not have a meaningful impact on smoking cessation.

An important motivation for studying the heterogeneous impact of tobacco excise on smoking behaviour was that the decrease in smoking consumption has stalled in the past 20 years (Orzechowski & Walker, 2016). Our findings suggest that excise taxes are an effective method to reduce tobacco usage, even among the group with a high genetic predisposition towards smoking. Even more, those with a high genetic predisposition to smoking respond most strongly to changes in tobacco excise taxes. However, the interaction between the genetic predisposition to smoking and tobacco excise taxes does not significantly impact smoking cessation in our sample. The insignificance of this interaction could be driven by the relatively small sample size in these analyses. However, it may also somewhat reflect the age composition of the sample. The HRS samples individuals aged over 50 years and their spouses, and several smokers in the sample may already have been smoking for the largest part of their life and the effect of further increases in tobacco excise taxes may no longer be dependent on their genetic predisposition. Nevertheless, tobacco taxes and the genetic predisposition for smoking cessation additively impact the decision to remain smoking. Thus, overall, our findings are largely in line with (Boardman, 2009) showing that environmental circumstances such as state policies (including taxation policies) moderate the effect of genes on smoking.

Although Fletcher (2012) was the first to show that only individuals carrying a certain genetic variant respond to increases in excise taxes, Fontana (2015) provided evidence that population stratification was driving these initial results. However, based on a weighted combination of multiple (approximately 1.4 million SNPs) genetic variants, i.e., a polygenic score, in the present study we do find again a significant interaction effect along the intensive and extensive margin for smoking. The sample restriction to individuals of European ancestry and the inclusion of principal components makes that the present findings are not likely to be driven by (subtle forms of) population stratification. However, in contrast with the findings of Fontana (2015), we do find a significant impact of the interaction between the genetic predisposition to heavy smoking and excise taxes on someone’s smoking behaviour. We believe this is the result of us using polygenic scores with higher predictive power which are also more closely related to the smoking outcomes analysed. Moreover, our analyses were also more powerful as the results of having a larger analysis sample both in terms of individuals and observations. As such, the present findings contribute to the literature analysing heterogeneity in smoking behaviour (Nesson, 2017) by highlighting genes as an important factor moderating the response to tobacco excise taxes.

Our *gene-environment interaction* study goes beyond the earlier *heritability-environment interaction* study (Boardman, 2009) by using individual-level molecular genetic information to analyse smoking behaviour. As such, it allows for analyses evaluating the effectiveness of policies on the individual-level (Benjamin et al., 2011; Manski, 2011). Although this does not imply that it is possible to accurately predict individual-level behavioural outcomes (Rietveld et al., 2020), from a policy perspective our findings clearly suggest that there is genetic heterogeneity in response to tobacco excise taxes. Individuals with a high genetic predisposition towards smoking respond stronger to tobacco excise taxes compared to individuals with a lower genetic predisposition. Thus, large increases in tobacco excise taxes lower smoking behaviour more among those with a high genetic predisposition for smoking than could expected based on the effect of excise taxes alone. Before starting to develop policies based on these findings, further research is needed to understand what exactly makes that those with a low genetic propensity for smoking respond relatively mildly to changes in tobacco excise taxes. The polygenic scores relate to biological systems that affect reward processing and addiction (Liu et al., 2019), and it may be that the nature of smoking behaviour (e.g., addiction vs. recreational use) differs between those with a high or low genetic predisposition for smoking. If so, although our findings do show that tobacco excise taxes are an effective policy instrument on their own, the present study suggests that different policies for genetically different types of individuals could be developed to further reduce smoking in the population.

Despite the significance of our finding, our study is not without limitations. Importantly, HRS participants are only surveyed every two years. In the analyses, we therefore used the excise taxes one year before each smoking measurement. This may be less suitable if the response time to increases in excise taxes differs among individuals. Also, individuals who live close to the border of a state could purchase their tobacco in the neighbouring state with a lower excise tax on tobacco (Chiou & Muehlegger, 2008). In our analyses, we cannot rule out whether this is driving our results. Another limitation of the current sample is that it is a representative sample of older Americans only. As over time only the most addicted individuals are likely to remain smoking, this set of individuals might be particularly insensitive to changes in excise taxes. Therefore, we consider the replication of the present results in a younger sample to be particularly relevant. Finally, our sample is restricted to individuals of recent European ancestry. Smoking behaviour is known to differ across ethnicities (Kandel et al., 2004), and as such we miss out an important source of heterogeneity in our analyses. To deal with population stratification bias, the restriction to one ancestry group is necessary but future studies may attempt to replicate the present findings in samples of other ancestries. However, as GWAS samples are relatively small for other ancestries, this may not be possible in the near future.

## Data Availability

HRS data are available (after registration) via https://hrsdata.isr.umich.edu/

## Acknowledgements

We thank the National Institute on Aging (U01 AG009740), the Health and Retirement Study (HRS application number HRS RDA 2019-025) staff, and the HRS participants. C.A.R. acknowledges funding from the New Opportunities for Research Funding Agency Cooperation in Europe (NORFACE-DIAL grant 462-16-100).

## Appendix

### Regressions in subsamples based on quartiles of the polygenic score distribution

In our main analyses presented in the main text, we focus on the multiplicative linear interaction between the polygenic scores and excise tobacco taxes. We find significant interactions in the models explaining an individual’s current smoking status and in the models explaining the intensity of smoking. In order to investigate possible non-linear interactions, we here provide regressions results obatained in subsamples based on quartiles of the distribution of the polygenic scores.

Table 5 present the results for the regressions explaining an individual’s current smoking status, and shows that the effect of tobacco excise taxes gradually increases over the quartiles, which suggests that the linear interaction term fits the data adequately. Table 6 present the 20 result of the regression explaining the amount of tobacco consumption. It shows that the effect of tobacco excise taxes increases over the four quartiles, but the increase in effect size flattens somewhat off in the fourth quartile.

**Table 5:** Results of the regressions explaining an individual’s current smoking status in subsamples based on quartiles of the polygenic score for smoking initiation.

|  | (1) | (2) | (3) | (4) |
| --- | --- | --- | --- | --- |
| -Log(Tax) | 0.042***<br>(0.007) | 0.064***<br>(0.008) | 0.077***<br>(0.007) | 0.083***<br>(0.007) |
| Female | -0.001<br>(0.007) | -0.024*<br>(0.009) | -0.021<br>(0.012) | -0.059***<br>(0.014) |
| Birth year | 0.221*<br>(0.102) | -0.033<br>(0.148) | 0.031<br>(0.122) | -0.060<br>(0.139) |
| Birth year<sup>2</sup> | -0.000*<br>(0.000) | 0.000<br>(0.000) | -0.000<br>(0.000) | 0.000<br>(0.000) |
| Income ($\times \$1,000$) | -0.000**<br>(0.000) | -0.000***<br>(0.000) | -0.000*<br>(0.000) | -0.000<br>(0.000) |
| Years of education | -0.011***<br>(0.002) | -0.016***<br>(0.003) | -0.016***<br>(0.002) | -0.023***<br>(0.002) |
| Married | -0.064***<br>(0.012) | -0.072***<br>(0.012) | -0.083***<br>(0.013) | -0.097***<br>(0.010) |
| Observations | 26,537 | 26,521 | 26,460 | 26,441 |
| Individuals | 3,016 | 3,005 | 3,015 | 3,022 |
| $R^2$ | 0.0467 | 0.0618 | 0.0678 | 0.0945 |

*Notes:* Standard errors in parentheses (clustered by state and individual); Coefficients for the constant term and the principal components are not reported, but available upon request from the authors;
\* $p < 0.05$ , \*\* $p < 0.01$ , \*\*\* $p < 0.001$ .

**Table 6:**
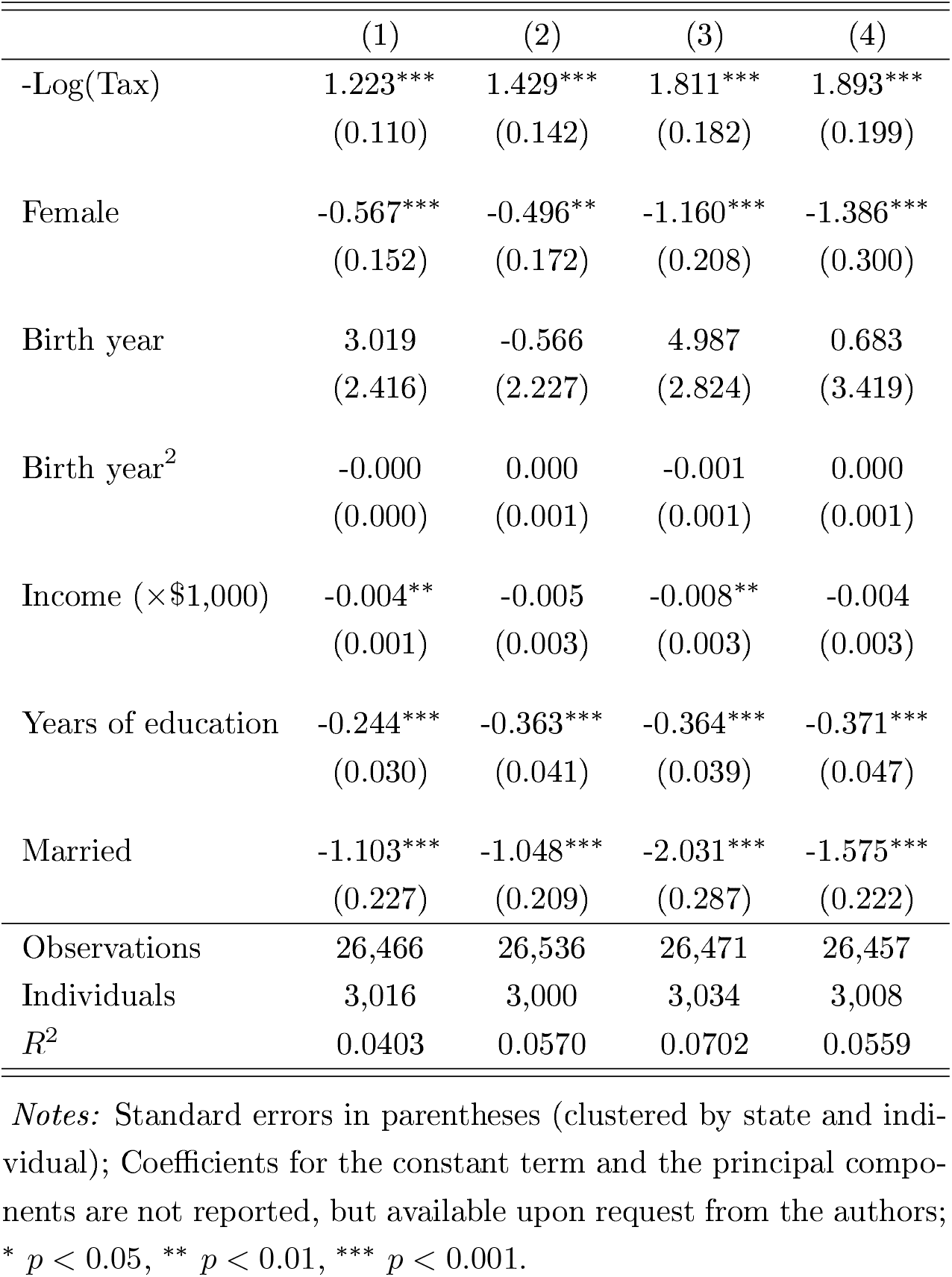
Results of the regressions explaining an individual’s smoking intensity in subsamples based on quartiles of the polygenic score for smoking intensity.

## Footnotes

1 When using logit specifications, we obtain qualitatively similar results. These results are available upon request from the authors.

2 To enhance the interpretation of the interaction effects, we multiply Tax_*st*_ in all regression with - 1. By doing so, we ensure that the effects for Tax_*st*_ as well as for the polygenic scores are expected to be positive.

3 In Table 5 in the Appendix, we provide regression results for quartiles of the sample based an individual’s value for the polygenic score. In line with the results presented here in the main text, the results also suggest a stronger response to lower tobacco excise taxes by individuals with higher values for the polygenic score.

4 Additional analyses, reported in Table 6 in the Appendix, also show that the response to tobacco excise taxes increases over the quartiles of the polygenic score distribution.

5 To let the polygenic score have a positive effect on the outcome variable, the dependent variable is coded as 1 for those continuing smoking and 0 for those that stopped smoking in these models. Note that PGS for smoking cessation also reflects the likelihood to remain smoking, see section 2.3.

## Notes

### Competing Interest Statement

The authors have declared no competing interest.

### Author Declarations

Erasmus Research Institute of Management (IRB NE 2019-15)

